# A functional hierarchy of semicircular canal and otolith contributions to early motor development

**DOI:** 10.64898/2026.05.08.26352737

**Authors:** Marta Campi, Sylvette R. Wiener-Vacher, Audrey Maudoux, Hung Thai-Van

## Abstract

The vestibular system is critical for early motor development, yet the respective roles of its two subsystems, the semicircular canals and otolith organs, remain poorly defined. Here we analyse 411 children with comprehensive vestibular assessment to determine whether a functional hierarchy underlies their contributions to the acquisition of four postural and motor milestones during the first two years of life. Using Type III ANOVA to account for the frequent co-occurrence of canal and otolith dysfunction, we show that canal areflexia is associated with a 7.0-month delay in independent walking, three times larger than the otolith effect. Canal function is the only component reaching significance after Bonferroni correction across four milestones. Canal function alone predicts walking delay (>18 months) with an area under the curve of 0.83. Canal areflexia carries a positive predictive value of 80.2% for walking delay, while normal canal function effectively rules out a walking delay of vestibular origin (negative predictive value 93.5%). These findings establish a functional hierarchy of vestibular contributions to motor development and identify canal function as a powerful developmental biomarker.

## Introduction

The acquisition of postural and motor control in early childhood relies on the integration of multiple sensory inputs, including visual, vestibular, somatosensory, and proprioceptive signals [1,2,9]. This process, often described as multisensory integration, enables the central nervous system to dynamically reweight sensory information to maintain balance and guide action [3,10,15]. During the first years of life, this progressive calibration supports the sequential acquisition of developmental milestones such as head control, sitting, standing, and independent walking.

Among these sensory systems, the vestibular system plays a critical role. Vestibular dysfunction, whether partial or complete, has been consistently associated with delays in key motor milestones, particularly when occurring during critical developmental periods [7,8,17,21,25,26]. Beyond motor delay, vestibular loss has also been linked to broader impairments in psychomotor development [3,22]. While these findings establish the importance of vestibular input as a whole, they do not resolve how its distinct components contribute to motor development.

The vestibular system comprises two functionally distinct subsystems: the semicircular canals, which encode angular head motion, and the otolith organs (saccular and utricular receptors), which encode linear acceleration and gravity. Their distinct encoding properties suggest that they may differentially support postural control and motor acquisition. Developmental studies indicate that both canal- and otolith-mediated responses undergo substantial maturation during early childhood, as reflected by changes in vestibulo-ocular reflex gain [11,19,20,23,24]. However, the respective contributions of these subsystems to the timing of specific developmental milestones remain poorly understood. In particular, it is unknown whether semicircular canal and otolith signals contribute equally to motor development, or whether a functional hierarchy exists between them. Addressing this question requires disentangling their independent and interactive effects across a broad spectrum of vestibular functions.

Here, we take advantage of a large cohort of 411 children undergoing comprehensive vestibular assessment to quantify the respective contributions of semicircular canal and otolith function to early motor development, and to evaluate the clinical predictive value of canal function for identifying children at risk of developmental delay. By analysing the principal postural and motor acquisition milestones during the first two years of life across a wide range of vestibular impairments, we demonstrate a hierarchical organisation of vestibular contributions, in which semicircular canal function emerges as the primary determinant of motor milestone timing, with otolith function playing a secondary role.

## Results

### Study population

Canal function (FC) and otolith function (FO) were each classified as normal (2), partial (1), or absent/areflexia (0), based on comprehensive vestibular testing including video head impulse test (vHIT), caloric and earth vertical axis rotation chair tests for canals, and cervical vestibular evoked myogenic potentials (cVEMP) for otoliths (see Methods and Supplementary Table 1 for detailed testing criteria).

A total of 411 children were included (205 females, 206 males; mean age at vestibular testing 20.2 months, SD 8.5, range 2.3-36.9 months; Figure 1). None had neurological disorders. Normal vestibular function (NVF: FO=2 and FC=2) was present in 163/411 children (39.7%) and complete bilateral vestibular loss (CBVL: FO=0 and FC=0) was present in 81/411 (19.7%). The remaining 167/411 (40.6%) had partial vestibular loss (PVL) with various FO and FC combinations. The distribution across FO and FC combinations was highly asymmetric: of 135 children with FO=0, 81/135 (60.0%) also had FC=0, and only 3 (2.2%) had FC=2 (Table 1). This co-occurrence underscores the importance of statistical adjustment when estimating each vestibular component’s independent contribution.

**Table 1.** Distribution of vestibular function status and age of acquisition of the postural and motor milestones for the 411 children. Nine FO×FC combinations shown with sample sizes and mean milestone ages. VF = vestibular function group; FO = otolith function (0=areflexia, 1=partial, 2=normal); FC = canal function. NVF = normal vestibular function (FO=2, FC=2); PVL = partial vestibular loss; CBVL = complete bilateral vestibular loss (FO=0, FC=0). Ages in months. SD = standard deviation.

| VF | FO | FC | FO+FC | N (%) | Head Holding | Sitting | Standing | Walking |
| --- | --- | --- | --- | --- | --- | --- | --- | --- |
|  |  |  |  |  | Mean (SD) | Mean (SD) | Mean (SD) | Mean (SD) |
| NVF | FO=2 | FC=2 | FO=2 + FC=2 | 163 (39.7) | 3.0 | 7.0 | 10.0 | 13.0 |
| NVF |  |  |  |  | 3.1 (1.1) | 7.0 (1.5) | 10.2 (2.0) | 14.0 (2.8) |
| PVL | FO=2 | FC=0 | FO=2 + FC=0 | 2 (0.5) | 6.0 | 10.5 | 12.0 | 22.0 |
| PVL |  |  |  |  | 6.0 (1.4) | 10.5 (0.7) | 12.0 (0.0) | 22.0 (2.8) |
| PVL | FO=2 | FC=1 | FO=2 + FC=1 | 75 (18.2) | 4.0 | 7.0 | 12.0 | 15.0 |
| PVL |  |  |  |  | 3.7 (1.6) | 7.4 (1.9) | 11.5 (2.2) | 15.1 (3.3) |
| PVL | FO=0 | FC=2 | FO=0 + FC=2 | 3 (0.7) | 4.2 | 6.5 | 12.0 | 16.5 |
| PVL |  |  |  |  | 4.2 (1.8) | 7.5 (2.2) | 12.0 (0.0) | 15.8 (1.6) |
| PVL | FO=1 | FC=2 | FO=1 + FC=2 | 3 (0.7) | 3.0 | 11.5 | 20.0 | 15.0 |
| PVL |  |  |  |  | 3.0 (0.0) | 11.5 (9.2) | 20.0 (0.0) | 17.0 (6.2) |
| PVL | FO=1 | FC=0 | FO=1 + FC=0 | 8 (1.9) | 5.0 | 9.5 | 15.0 | 23.5 |
| PVL |  |  |  |  | 4.3 (2.1) | 11.6 (4.9) | 15.2 (3.0) | 23.4 (3.7) |
| PVL | FO=1 | FC=1 | FO=1 + FC=1 | 25 (6.1) | 3.0 | 9.0 | 12.0 | 18.0 |
| PVL |  |  |  |  | 3.6 (1.1) | 8.8 (1.7) | 12.2 (3.0) | 17.9 (2.9) |
| PVL | FO=0 | FC=1 | FO=0 + FC=1 | 51 (12.4) | 4.5 | 9.0 | 12.0 | 18.0 |
| PVL |  |  |  |  | 5.2 (2.2) | 9.1 (2.5) | 13.1 (3.9) | 19.3 (4.3) |
| CBVL | FO=0 | FC=0 | FO=0 + FC=0 | 81 (19.7) | 5.2 | 10.0 | 14.8 | 22.0 |
| CBVL |  |  |  |  | 5.8 (2.7) | 10.6 (3.0) | 14.5 (3.1) | 22.5 (4.4) |

**Figure 1.**
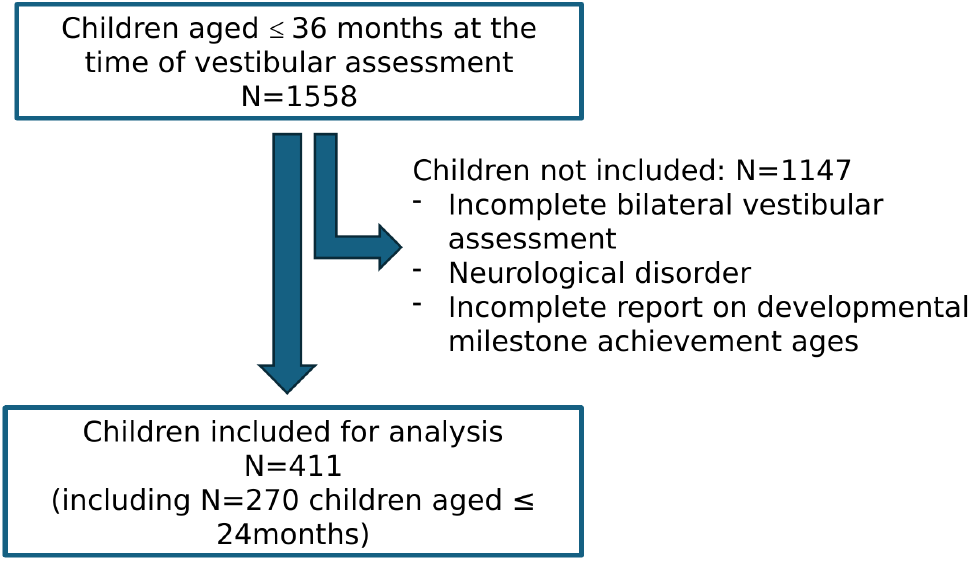
Flow diagram of study population. Children aged up to 36 months at the time of vestibular assessment (N=1,558). After exclusion of children with incomplete bilateral vestibular assessment, neurological disorder, or incomplete milestone data, N=411 children were included for analysis (including N=270 children aged ≤24 months).

The distribution of acquisition ages for each milestone are shown in Figure 2 and compared to the data from a healthy paediatric population [18]. The median ages of milestone acquisition across all nine FO and FC combinations are depicted in Figure 3, which highlights isolated otolith loss (red boxes, FO=0 and FC=2, n=3) and isolated canal loss (blue boxes, FO=2 and FC=0, n=2). For each milestone, the median age of acquisition was significantly higher for children with PVL and CBVL compared to NVF (Kruskal-Wallis test, all p<0.001; Table 2).

**Table 2.** Median ages of acquisition of each postural and motor control milestone in children with NVF (n=163), PVL (n=167), and CBVL (n=81). Kruskal-Wallis test p-values and Dunn’s pairwise comparisons with Bonferroni correction. Ages in months.

| Milestone | Vestibular function | N | Median age of acquisition | 95%CI | P value |
| --- | --- | --- | --- | --- | --- |
| Head Holding | NVF | 61 | 3 | [1; 5] | — |
| Head Holding | PVL | 82 | 4 | [2; 9] | < 0.0001 |
| Head Holding | CBVL | 54 | 5.2 | [2.3; 12] | < 0.0001 |
| Sitting | NVF | 93 | 7 | [4.3; 9] | — |
| Sitting | PVL | 118 | 8 | [5; 16.1] | < 0.0001 |
| Sitting | CBVL | 70 | 10 | [5; 18] | < 0.0001 |
| Standing with support | NVF | 76 | 10 | [6.9; 15] | — |
| Standing with support | PVL | 80 | 12 | [8; 20] | < 0.0001 |
| Standing with support | CBVL | 46 | 14.8 | [9; 19.9] | < 0.0001 |
| Independent walking | NVF | 162 | 13 | [10; 21.9] | — |
| Independent walking | PVL | 167 | 17 | [11; 26] | < 0.0001 |
| Independent walking | CBVL | 81 | 22 | [15; 32] | < 0.0001 |

**Figure 2.**
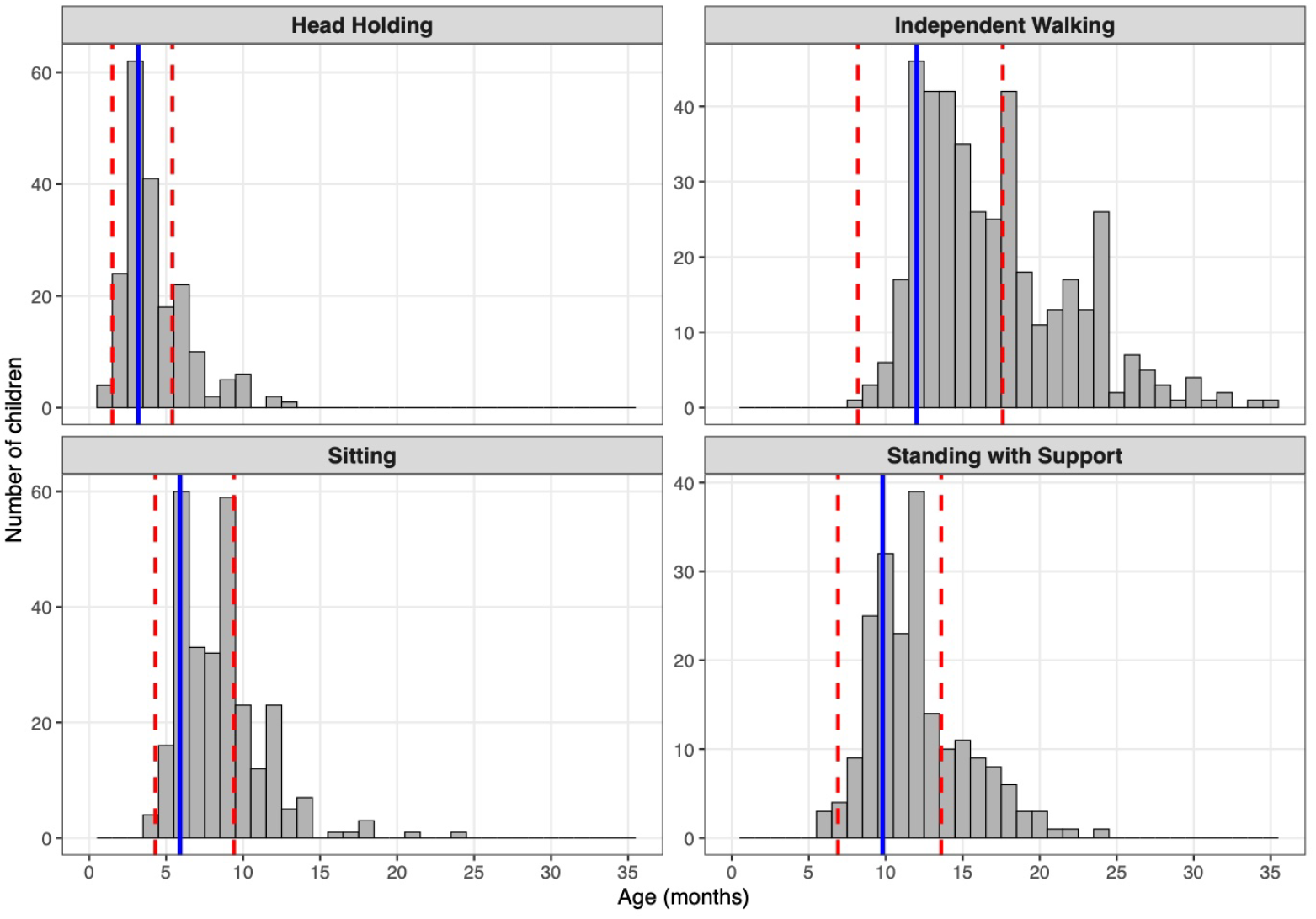
Distribution of milestone acquisition ages compared to WHO reference. Distribution of acquisition ages for each of the four milestones in the 411 children included in the study compared to reference values from the WHO Multicentre Growth Reference Study, 2006-2008. Blue solid line: WHO median age of acquisition. Red dashed lines: WHO 5th and 95th percentiles. Note that for the included children, the mean age of milestone acquisition is higher than the reference values from the healthy paediatric population.

**Figure 3.**
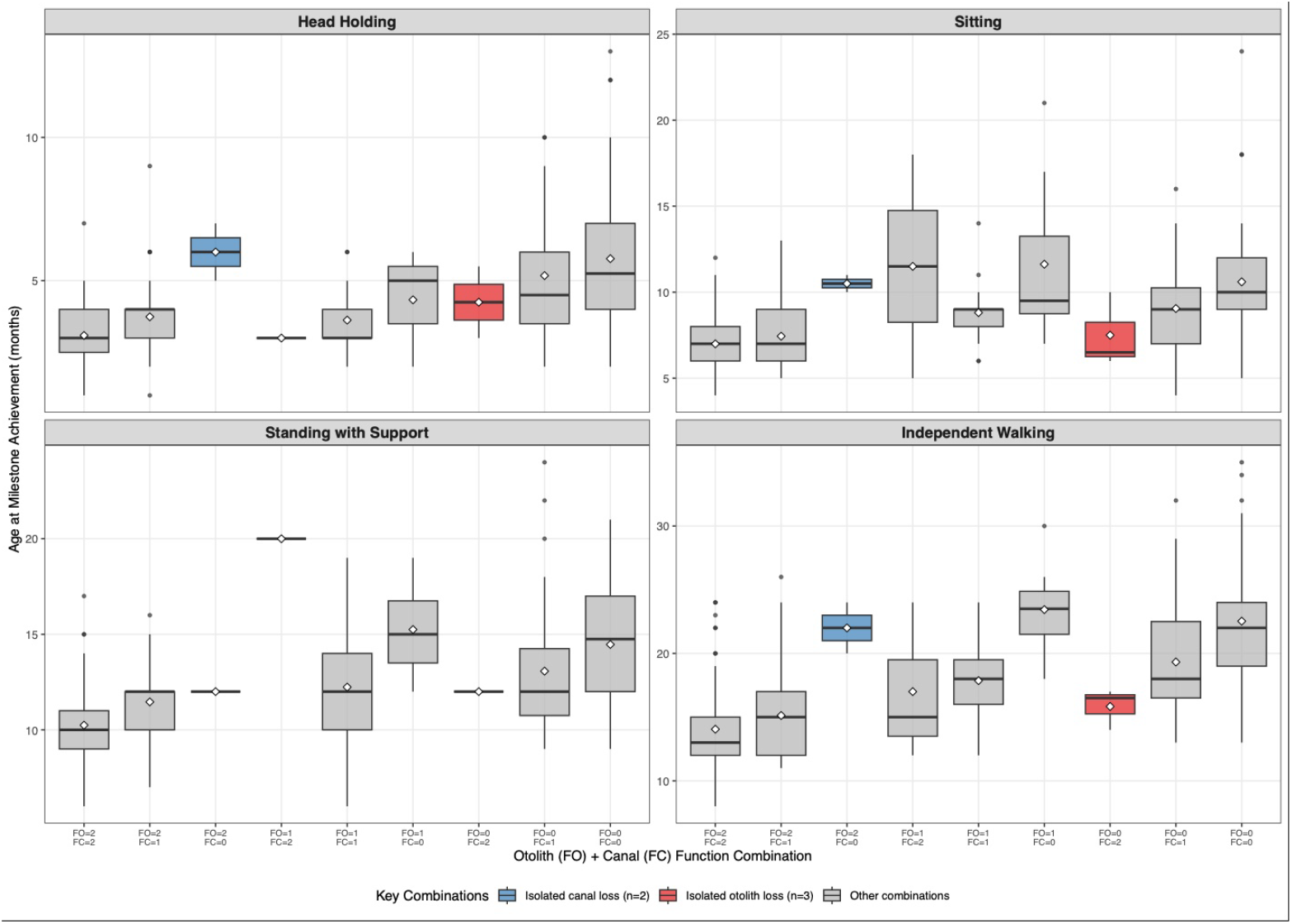
Distribution of milestone achievement ages by vestibular function combination. Boxplots show age distribution (months) at which each milestone was achieved, organised by FO×FC combination. Panels are ordered by developmental sequence: head holding, sitting, standing with support, and independent walking. Red boxes: isolated otolith loss (FO=0, FC=2; n=3). Blue boxes: isolated canal loss (FO=2, FC=0; n=2). Grey boxes: all other combinations. White diamonds = group means; box = interquartile range (25th—75th percentile); whiskers = extend to 1. 5× IQR; individual points = outliers beyond whiskers. FO = otolith function; FC = canal function (0 = areflexia, 1 = partial, 2 = normal). Note: isolated loss groups (n=2–3) are highlighted for visual reference but small sample sizes limit statistical interpretation.

### Canal function is the primary determinant of milestone acquisition timing

Type III two-way ANOVA showed that FC had the largest effect on milestone timing (Table 3, Figure 4A). All p-values reported below are uncorrected; the Bonferroni-corrected threshold for significance across four milestones is ×= 0.05/4 = 0.0125. Three milestones (head holding, sitting, independent walking) showed no significant FC×FO interaction (all p>0.25), allowing interpretation of independent main effects. Standing with support showed a significant interaction (p=0.037), requiring specific FO×FC combination effects to be examined (Figure 4B).

**Table 3.** Canal and otolith contributions to developmental milestone timing (Type III ANOVA). Type III p-values for FO, FC, and FO×FC interaction; adjusted marginal mean delays with 95% CIs; partial 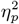; canal-to-otolith delay ratio. For milestones without significant interaction main effects are reported (see Figure 4A); for the milestone with significant interaction (standing with support) interaction effects are reported (see Figure 4B). FO = otolith function; FC = canal function. 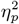= partial eta-squared.

| Milestone | n | FO×FC p | Canal delay (mo) | [95% CI] | Canal p | Canal $\eta_p^2$ | Otolith delay (mo) | [95% CI] | Otolith p | Otolith $\eta_p^2$ | Ratio |
| --- | --- | --- | --- | --- | --- | --- | --- | --- | --- | --- | --- |
| Head Holding | 197 | 0.863 | 1.9 | [-0.0, 3.9] | 0.053 | 0.024 | 0.8 | [-0.5, 2.1] | 0.244 | 0.015 | 2.44 |
| Sitting | 281 | 0.297 | 2.2 | [0.3, 4.2] | 0.022 | 0.046 | 0.7 | [-0.8, 2.2] | 0.333 | 0.026 | 3.04 |
| Standing with Support | 202 | 0.037 | - | See Fig. 4B | - | 0.02 | - | See Fig. 4B | - | 0.053 | - |
| Independent Walking | 410 | 0.307 | 7 | [4.4, 9.7] | <0.001 | 0.077 | 2.2 | [0.0, 4.3] | 0.049 | 0.013 | 3.24 |

**Figure 4.**
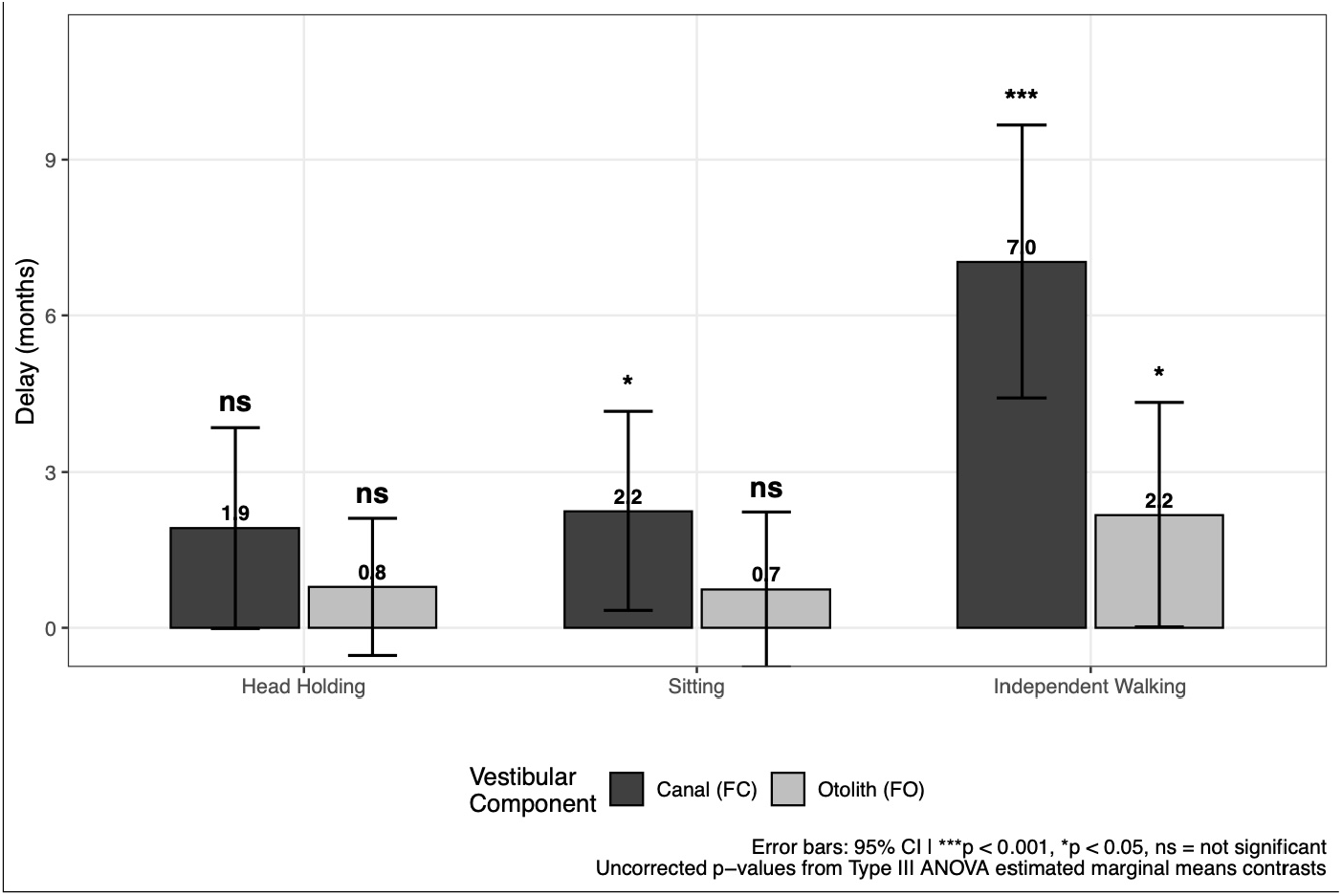
(A) Developmental delays for milestones without FC×FO interaction (Type III ANOVA). Adjusted mean delays (months) with 95% confidence intervals for canal areflexia (FC=0, dark grey) and otolith are-flexia (FO=0, light grey) compared to normal function (FC=2 or FO=2, respectively). Each effect is statistically controlled for the other vestibular component using Type III sums of squares (**p<0. 001, *p<0. 05, ns = not signif-icant). Head holding, sitting, and independent walking milestones are shown here as there is no significant FC×FO interaction for these milestones (all p>0. 25). Standing with support is not represented because there is a significant FC×FO interaction for this milestone (p=0. 037; see Figure 4B).

**Figure 4.**
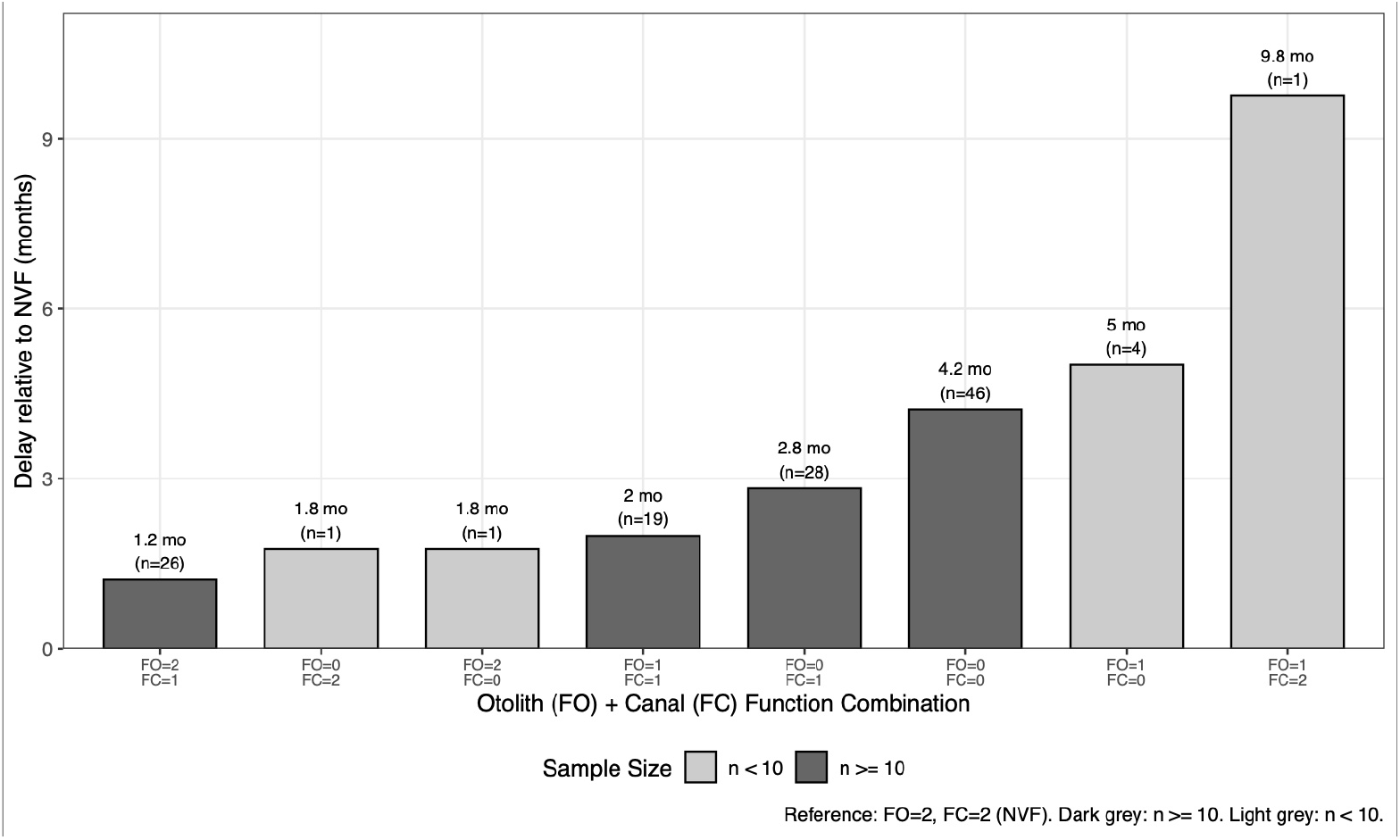
(B) Standing with support: combination effects due to FC×FO interaction. Significant FC×FO interaction (p=0. 037) indicates that canal and otolith effects cannot be interpreted independently for this milestone. Graph shows mean delays (months) compared to normal function baseline (FO=2, FC=2; NVF, n=76) for all nine FO×FC combinations. Values above bars indicate mean delay in months and sample size. Dark grey bars: adequate sample size (n≥10) for reliable interpretation. Light grey bars: small sample size (n<10) requiring cautious interpretation due to limited statistical power. The interaction pattern suggests that for standing, intact canal function may partially compensate for otolith dysfunction, and vice versa, whereas for walking, sitting, and head holding (Figure 4A), the two systems contribute more independently.

The adjusted mean delays estimated for head holding were 1.9 months for FC=0 (95% CI: −0.0-3.9, p=0.053, partial 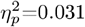, small effect) and 0.8 months for FO=0 (95% CI: −0.5-2.1, p=0.244, partial 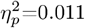, small effect). Neither reached significance at the uncorrected level (both p>0.05), likely reflecting the substantial missing data for this early milestone (52% missing). The canal-to-otolith delay ratio was 2.4 months (Table 3).

Regarding sitting, FC=0 was associated with a 2.2-month adjusted mean delay (95% CI: 0.3-4.2, p=0.022, partial 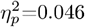, small effect). This effect was significant at the uncorrected level (*α* = 0.05) but did not reach the Bonferroni-corrected threshold (*α*=0.0125). FO=0 showed a non-significant 0.7-month adjusted mean delay (95% CI: −0.8-2.2, p=0.333, partial 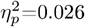, small effect). The canal-to-otolith delay ratio was 3.0 (Table 3).

Independent walking showed the strongest canal effect. FC=0 was associated with a 7.0-month adjusted mean delay compared to FC=2 (95% CI: 4.4-9.7, p<0.001, partial 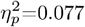, small-to-medium effect). FO=0 showed a 2.2-month adjusted mean delay (95% CI: 0.0-4.3, p=0.049, partial 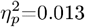, small effect). The canal-to-otolith delay ratio was 3.2 months (Table 3). After Bonferroni correction for four milestones (*α* = 0.05/4 = 0.0125), the canal effect remained significant (p<0.001) while the otolith effect did not (p=0.049).

Across all milestones without interaction, canal dysfunction consistently showed 2.4-to 3.2-fold greater adjusted mean delays than otolith dysfunction (Table 3, Figure 4A). FC was the only component reaching significance after correction for multiple comparisons. For independent walking, the otolith effect was significant at the uncorrected level (p=0.049) but did not reach the Bonferroni-corrected threshold (*α*=0.0125), consistent with a modest contribution substantially smaller than the canal effect.

Standing with support showed a significant FC×FO interaction (p=0.037, tested separately for this milestone), indicating that the effect of one vestibular component depends on the status of the other (Figure 4B). Because of this interaction, main effects cannot be meaningfully interpreted; instead, specific FO×FC combination effects were examined.

Among adequately powered combinations (n≥10), delays relative to NVF (FO=2 and FC=2, n=76, mean age 10.2 months) ranged from 1.2 months (FO=2 and FC=1, n=26) to 4.2 months (FO=0 and FC=0, n=46). The interaction pattern suggests that for standing, intact FC may partially compensate for otolith dysfunction, and vice versa, whereas for the other three milestones (Figure 4A), the two systems contribute more independently. Of note, small sample combinations (n=1-4) are shown in Figure 4B for completeness but should be interpreted with caution.

### Covariate adjustment and graded dysfunction

Canal effects remained significant after adjustment for sex and age at vestibular testing (walking: p<0.001; sitting: p<0.001; both surviving Bonferroni correction). Otolith function did not reach significance for walking after adjustment (p=0.115) and for sitting (p=0.052). Sex was a significant covariate for walking (p=0.021) and sitting (p=0.002, uncorrected exploratory tests), consistent with previously reported sex differences in the timing of acquisition of these milestones. Sex was not significant for standing or head holding, possibly reflecting the earlier developmental timing and higher proportion of missing data (51-52%) for these milestones. Age at testing was not significant for any milestone (Supplementary Table 2).

Linear regression analysis treating vestibular function levels as continuous (0=areflexia, 1=partial, 2=normal) confirmed that delays increase progressively with dysfunction severity for all four milestones (Supplementary Table 3). The strongest graded effect was observed for independent walking: each level of increasing canal dysfunction was associated with a 3.2-month increase in walking age (p<0.001, R^2^=0.48, indicating that vestibular function level explained 48% of the variance in walking age).

### Validation

In children ≤24 months at vestibular testing (n=270, 65.7% of sample), the canal effect remained the larger of the two vestibular contributions, consistent with the full sample (walking canal delay: p<0.01; otolith: not significant). Restricting FO×FC combinations to n≥10 (n=395) confirmed the findings of the primary analysis, demonstrating that results were not driven by combinations with small sample sizes. Permutation tests (10,000 permutations) confirmed the canal main effect for walking (permutation p<0.001) and sitting (permutation p<0.01). Bootstrap 95%CIs (5,000 BCa resamples) for the canal-to-otolith delay ratio excluded 1.0 for walking and sitting (Supplementary Tables 4-7). Model diagnostics revealed non-normal residuals for all milestones (Shapiro-Wilk, all p<0.001), but permutation test results confirmed that parametric findings were robust to this departure (Supplementary Table 8).

### Canal function predicts independent walking onset delay

To evaluate whether vestibular assessment can be used for clinical risk stratification, we fitted logistic regression models predicting developmental delay, defined as independent walking onset >18 months (see Methods). Developmental delay (independent walking onset >18 months) occurred in 129/411 children (31.4%). Delay rates were 80.2% for FC=0 (n=91), 29.8% for FC=1 (n=151), and 6.5% for FC=2 (n=168).

Logistic regression using FC (coded as 0, 1, 2) as the sole predictor achieved an AUC of 0.83 (95% CI: 0.79-0.87). Adding FO improved discrimination modestly but significantly (AUC=0.86, ΔAUC=0.03, likelihood ratio test p<0.001; DeLong test p<0.01). The full model including sex and age at vestibular testing achieved an AUC of 0.87. Ten-fold cross-validation confirmed these estimates (canal only: CV-AUC=0.80; canal+otolith: CV-AUC=0.83; Table 4, Figure 5; Supplementary Table 9).

**Table 4.** Logistic regression models predicting developmental delay (independent walking onset >18 months, 129/411 children = 31. 4%). Four nested models with AIC, AUC with 95% DeLong CIs, 10-fold CV-AUC, and likelihood ratio tests.

| Model | AIC | AUC [95% CI] | CV-AUC | ΔAUC vs Canal |
| --- | --- | --- | --- | --- |
| Canal only (FC) | 361.7 | 0.832 [0.79, 0.87] | 0.796 | — |
| Canal + Otolith (FC+FO) | 347.1 | 0.860 [0.82, 0.90] | 0.831 | +0.028, $p < 0.001$ |
| Full (FC×FO + Sex + Age) | 348.4 | 0.869 [0.83, 0.91] | 0.852 | +0.037 |

**Figure 5.**
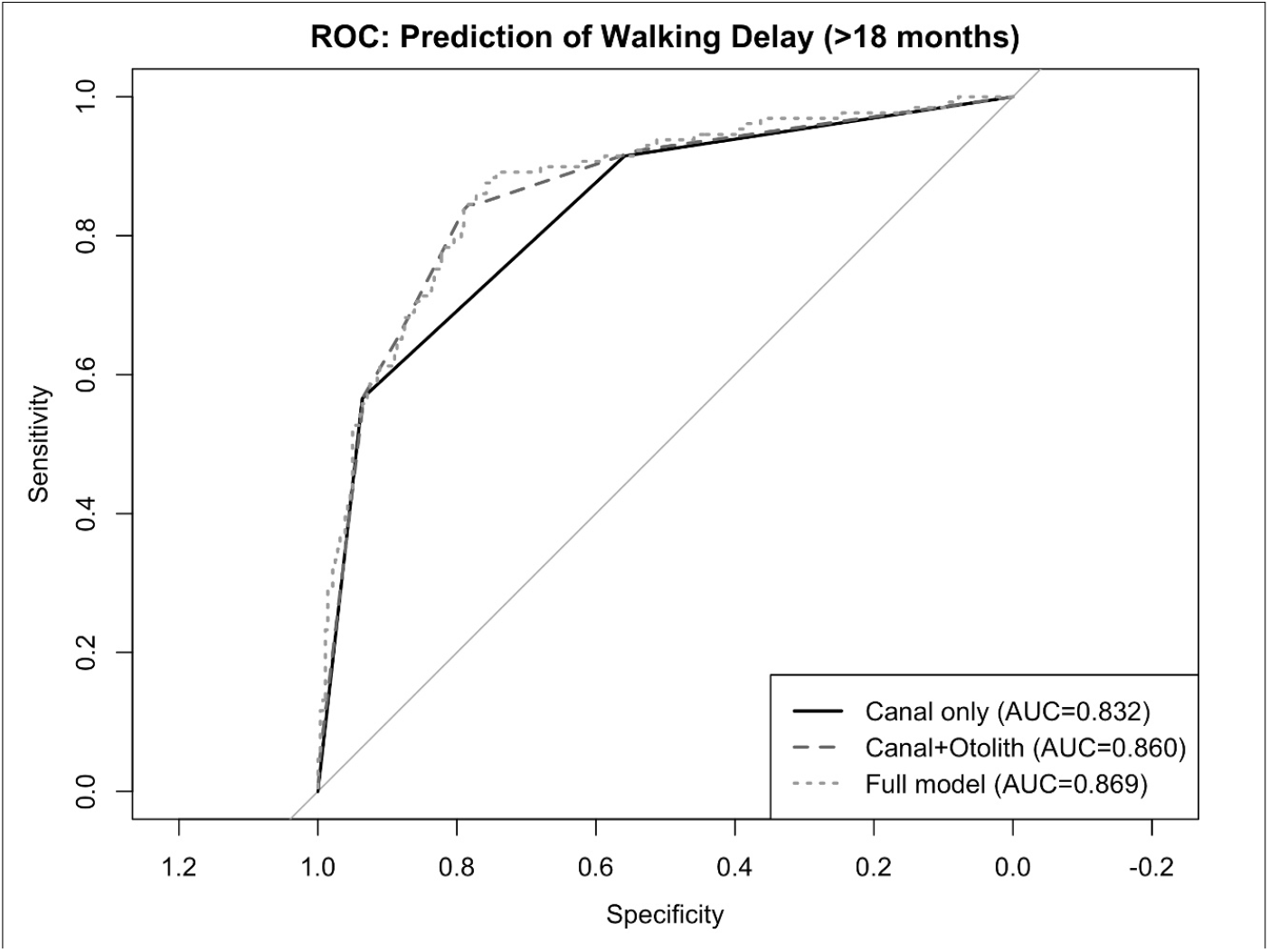
Receiver operating characteristic curves for prediction of walking delay (>18 months. ROC curves for three logistic regression models: canal function only (AUC=0. 83), canal + otolith function (AUC=0. 86), and full model including sex and age at testing (AUC=0. 87). DeLong test for canal vs canal+otolith: p<0. 01.

When using FC=0 as a clinical decision rule, a high specificity (93.6%) and positive predictive value (PPV, 80.2%) with moderate sensitivity (56.6%) for developmental delay was observed. FC=2 effectively ruled out developmental delay (negative predictive value (NPV) of 93.5%). Notably, developmental delay in the presence of FC=2 was rare: among children with PVL but normal FC (FC=2), only 1.2% had walking delay >18 months. Combined with the low delay rate in the NVF group (5.4%), this indicates that developmental delay in the presence of normal FC is unlikely to be vestibular in origin, supporting the high NPV and suggesting that alternative causes such as neurological disorders should be investigated in such cases. Any canal dysfunction (FC≤1) as a screening criterion captured 91.5% of children with developmental delay (Table 5).

**Table 5.** Clinical decision rules for walking delay (>18 months) based on canal function. Sensitivity, specificity, PPV, and NPV for three rules.

| Clinical Rule | Sensitivity | Specificity | PPV | NPV |
| --- | --- | --- | --- | --- |
| FC=0 → walking delayed | 56.6% (73/129) | 93.6% (263/281) | 80.2% | 82.4% |
| FC=2 → walking not delayed | — | — | — | 93.5% |
| FC≤1 → screening (at risk) | 91.5% (118/129) | 55.9% (157/281) | 48.8% | 93.5% |

## Discussion

### Main findings

This study provides quantitative evidence that semicircular canal function is the primary vestibular determinant of postural and motor milestone acquisition in early childhood. After accounting for the strong co-occurrence of canal and otolith dysfunction using Type III ANOVA, canal areflexia was associated with a substantial delay in independent walking (+7.0 months), an effect more than threefold larger than that observed for otolith areflexia. Canal function was the only vestibular component reaching significance after correction for multiple comparisons across all milestones.

Although smaller in magnitude, otolith function contributed modestly to predictive models indicating a secondary but measurable role. Canal function alone showed good discriminative performance for predicting delayed walking (>18 months), supporting its relevance as a developmental biomarker.

### A hierarchical organisation of vestibular contributions

These findings support the existence of a functional hierarchy within the vestibular system, in which semicircular canal signals act as the primary driver of motor milestone acquisition, while otolith signals provide a secondary contribution.

The dominance of canal function likely reflects its central role in dynamic balance control and gaze stabilisation through the vestibulo-ocular reflex, which are critical for coordinating head-body movements during emerging motor behaviours. In contrast, otolith-mediated encoding of gravity and linear acceleration may be partially compensated by other antigravitational sensory systems, including skin stretch receptors, muscle spindles, and Golgi tendon organs [6], which together provide redundant information about body orientation relative to gravity.

This distinction suggests two different modes of compensation: immediate redundancy for otolith-related functions via multisensory reweighting, versus delayed and training-dependent substitution for canal loss. While compensatory strategies can develop after canal loss, specifically catch-up saccades driven by the cerebellum and triggered by vision, these mechanisms do not exist under normal conditions and must be actively trained and maintained over time. This difference in compensatory availability may explain the substantially larger developmental impact of canal dysfunction.

Importantly, the findings herein indicate canal dominance but not otolith irrelevance: otoliths contribute meaningfully to developmental timing as a second-order factor, and this contribution may be underestimated by cVEMP’s assessment of only a restricted subset of the otolith receptor population [5]. It was previously shown that during the period of walking acquisition, otolith receptors (utricular and saccular receptors) could be functionally involved at different times of this critical developmental period [11].

### Developmental dynamics across milestones

The influence of canal function increased across the developmental sequence, with the largest effect observed for independent walking and significant effects also present for sitting. The weaker effect observed for head holding likely reflects limited statistical power due to missing data rather than a true absence of canal contribution.

A significant canal-otolith interaction was observed for standing with support, suggesting that this transitional stage may uniquely depend on the integration of dynamic (canal) and static (otolith) signals under specific biomechanical constraints. The interaction pattern indicates that for standing, intact canal function may partially compensate for otolith dysfunction, and vice versa, whereas for the other milestones, the two systems contribute more independently.

The role of canal function in predicting developmental milestone delays is consistent with previous reports of vestibular dysfunction affecting motor milestones; previous studies however, did not specifically disentangle canal and otolith contributions to developmental delay as vestibular loss was assessed before otolith testing became clinically available and thus only concerned canal function assessment [7,8,17,21,25]. The enhanced statistical analysis carried out herein confirms that canal function is the dominant contributor and the only component reaching significance after correction for multiple comparisons, while otolith function provides a smaller, supplementary contribution detectable in prediction models.

### Methodological considerations and interpretation of the otolith effect

The modest independent contribution of otolith function reflects both biological and methodological factors. The strong co-occurrence of canal and otolith dysfunction in this clinical population (60% of children with otolith areflexia also had canal areflexia) necessitated statistical deconvolution, which revealed a substantially smaller otolith effect after adjustment.

In addition, otolith function was assessed using cVEMP, which probes only a restricted subset of otolith receptors, specifically irregular, phasic type I cells of the striola, rather than the full macular hair cell population [5,13]. Furthermore, studies using off-vertical axis rotation (OVAR) have shown that saccular and utricular otolith responses mature differentially during early childhood, particularly around the age of independent walking acquisition [19,20]. This contrasts with the broader assessment of canal function using three complementary tests (vHIT, caloric, rotatory chair). The apparent limited contribution of otolith function may therefore partly reflect measurement constraints. Future development of more comprehensive otolith assessment methods (e.g., measurement of ocular torsion during head tilt) will be necessary to determine whether the observed hierarchy making the canal dominant reflects true physiological organisation or is influenced by current testing limitations.

### Clinical implications

These findings have direct implications for clinical practice, particularly in populations at high risk of vestibular impairment such as children with sensorineural hearing loss.

Canal function alone provides substantial predictive information for motor delay, with high NPV when normal (93.5%) and strong risk stratification when absent (80.2% of children with canal areflexia had delayed walking). Notably, developmental delay in children with normal canal function was rare even among those with PVL (1.2%), indicating that such delays are unlikely to be vestibular in origin and should prompt neurological evaluation. This supports a canal-first screening strategy using vHIT, which is rapid and increasingly accessible in paediatric settings [14].

For children with sensorineural hearing loss or suspected vestibular dysfunction, our findings suggest risk stratification based on canal function assessed in early infancy: canal areflexia predicts significant delays across all four developmental milestones; partial canal dysfunction is associated with moderate delays; and normal canal function effectively rules out vestibular origin of any observed developmental delay, prompting evaluation for neurological or other causes. Screening programmes for vestibular loss in children with profound hearing loss first proposed cVEMP testing from 6 months of age [12]. The present findings however, suggest that canal assessment (vHIT) should be prioritised as the primary screening tool, as canal function alone captures the majority of predictive information for developmental delay. cVEMP testing however, may be useful in case of canal dysfunction, despite its modest predictive value contribution in the model herein.

### Study limitations

Several limitations should be considered. The retrospective design precludes analysis of individual developmental trajectories. The rarity of isolated vestibular subsystem loss (only 3 children with isolated otolith loss and 2 with isolated canal loss) reflects the biological coupling of these structures but limits direct causal dissociation. This biological reality necessitates statistical deconvolution to isolate independent effects, which we achieved through Type III ANOVA. The high co-occurrence strengthens rather than weakens our approach: the canal effect remains dominant even when statistically controlling for this substantial overlap, and robustness analyses excluding combinations with small sample sizes confirmed all primary findings.

Missing data for head holding (52%) and standing (51%) reduced statistical power for these early milestones. This high proportion of missing data likely reflects that many children were tested after these milestones had already been acquired, with retrospective documentation being incomplete. In particular, head postural control was not described in detail in clinical reports and could not be reconstructed retrospectively. This pattern suggests that missingness is related to the age at clinical referral rather than to vestibular status or disease severity, reducing the risk of systematic bias. Potential confounders, including gestational age, socioeconomic factors, and family engagement were not controlled for. Finally, otolith function was assessed using cVEMP only, which principally tests saccular function. oVEMPs were not routinely performed during the period of the study due to the difficulty of obtaining reliable responses in young children. The otolith contribution reported here therefore reflects only saccular function limited to the striola receptors and may underestimate the full otolith contribution to motor development. External validation in an independent cohort is needed to confirm generalisability.

## Conclusion

Semicircular canal function is the primary vestibular determinant of early motor milestone acquisition, with effects substantially larger than those of otolith function after accounting for their co-occurrence. Canal are-flexia predicts walking delay with an AUC of 0.83, and normal canal function effectively rules out vestibularorigin for such delay. Notably, developmental delay in children with normal canal function was rare even among those with partial vestibular loss, indicating that such delays are unlikely to be vestibular in origin and should prompt neurological evaluation. These findings establish a hierarchical organisation of vestibular contributions to motor development, identify canal function as a robust biomarker for predicting developmental delay, and support prioritising canal assessment for early risk stratification and targeted intervention in children with suspected vestibular dysfunction.

## Methods

### Study design and population

This retrospective single-centre study analysed data collected over a 10-year period (2012-2023) in children having undergone a complete bilateral vestibular assessment when aged ≤36 months. To be included, children also had to have a description of developmental milestone acquisition ages, no history of neurological disorder, and normal clinical oto-neurological examination. Consent waivers were completed by parents or legal caregivers and were approved by an ethics committee (CPP-Ouest 3, number 2018-A01705-50). Independent walking age was available for 410/411 children; one child with normal vestibular function had no recorded walking age and was excluded from walking-specific analyses.

### Data collection and vestibular assessment

Sex, age at vestibular assessment, vestibular testing results, and ages of acquisition of four major developmental milestones (head holding, sitting, standing with support, independent walking) were collected from medical reports.

Complete bilateral vestibular assessment consisted in comprehensive clinical neuro-otological examination and instrumental vestibular testing covering a broad spectrum of vestibular sensitivity. For the purpose of this study, a complete vestibular assessment had to include these four tests. FC was tested using vHIT [23], bithermal caloric test [25], and earth vertical axis rotation chair test [19,20]. FO was tested using cVEMP. The criteria for normal and abnormal results to each test are presented in Supplementary Table 1.

### Vestibular function classification

FC was classified as: normal (FC=2) if responses to all tests (vHIT, caloric test, rotatory chair) were normal; partial (FC=1) if at least one test showed abnormal responses; absent (FC=0, canal areflexia) if no response was elicited in any of the tests. FO was classified as: normal (FO=2) if cVEMP responses were normal; partial (FO=1) if cVEMP responses were abnormal; absent (FO=0, otolith areflexia) if no cVEMP response was elicited. This scoring produced nine possible FO and FC combinations, forming a 3×3 matrix. NVF was defined as FO=2 and FC=2. CBVL was defined as FO=0 and FC=0. PVL comprised the seven remaining combinations (Table 1).

### Statistical analysis

All analyses were conducted in R version 4.2.1 (R packages listed below), with statistical significance set at *α*=0.05 for all tests.

#### Population characteristics

The Kruskal-Wallis rank-sum test was used to provide an initial descriptive comparison of median ages of milestone acquisition across the three vestibular function groups (NVF, PVL, CBVL), with Dunn’s test and Bonferroni correction for pairwise comparisons.

#### Primary analysis

To isolate the independent contribution of each vestibular component to developmental milestone timing, we performed two-way ANOVA for each milestone using the model: Age at milestone = FO + FC + FO×FC, where FO and FC are both treated as categorical factors with three levels (0=areflexia, 1=partial, 2=normal). Because the distribution across the nine FO and FC combinations was highly unbalanced (ranging from n=2 to n=163; Table 1), we used Type III sums of squares (`car` package, v3.1). This is necessary because 60% of children with FO=0 also had FC=0; without this adjustment, the canal and otolith effects would be confounded.

For milestones without significant interaction (p>0.05), the main effects were estimated using marginal means extracted from the `emmeans` R package (v1.8.0). These represent the mean age at milestone acquisition for each vestibular component function level (e.g., canal areflexia), averaged across all levels of the other component (e.g., otolith function), thereby statistically removing the influence of the other vestibular component. The adjusted mean delay was then calculated as the difference in estimated marginal means between areflexia (level 0) and normal function (level 2) for each component.

For milestones with significant interaction (p<0.05), main effects cannot be meaningfully interpreted since the effect of one vestibular component differs depending on the status of the other. For example, if FC=0 causes a larger delay when FO=0 or FO=1 compared to when FO=2, reporting a single canal effect averaged across otolith levels would be misleading. In such cases, specific FO and FC combination effects were examined instead.

Then, partial eta-squared 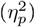 was used to quantify the proportion of variance in milestone acquisition timing that is explained by each vestibular component (i.e., how much of the variability when children reach a milestone can be attributed to FC or FO). Following Cohen [4], partial 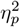 values of 0.01, 0.06, and 0.14 correspond to small, medium, and large effects. Cohen’s d was computed to express the difference between function levels (e.g., areflexia vs normal) in standard deviation units. Finally, the ratio of canal to otolith adjusted mean delays was calculated.

Bonferroni correction was applied for multiple comparisons across four developmental milestones (*α* = 0.05/4 = 0.0125 per milestone). Confidence intervals for estimated marginal means were calculated using the ANOVA model standard errors.

#### Covariate adjustment

Because sex and age at vestibular testing could independently influence milestone acquisition age (e.g., boys tend to walk later than girls), ANOVA models were re-estimated including these as additional covariates to verify that the observed canal and otolith effects are not driven by differences in sex or age at testing.

#### Graded dysfunction analysis

To test whether delays increase progressively with dysfunction severity (i.e., whether partial dysfunction causes intermediate delays between normal function and areflexia), linear regression was fitted for each milestone with age at milestone acquisition as the response variable and FO and FC treated as continuous predictors (coded as 0=areflexia, 1=partial, 2=normal) rather than categorical factors.

#### Validation

Three complementary approaches were carried out to validate the primary ANOVA findings (canal and otolith effects on milestone timing): (1) age-stratified analysis restricted to children ≤24 months at vestibular testing (n=270); (2) robust sample analysis restricted to FO×FC combinations with n≥10 children (n=395); and (3) non-parametric sensitivity analyses using permutation tests (10,000 permutations of FO and FC labels) and bootstrap confidence intervals (5,000 bias-corrected and accelerated [BCa] resamples) for adjusted mean delays and canal-to-otolith ratios.

#### Clinical prediction

To evaluate the clinical utility of vestibular assessment for predicting developmental delay, we focused on independent walking because it has a well-defined clinical threshold (>18 months), showed the largest canal effect (7.0 months), and the most complete data across milestone acquisition ages. Clinically delayed walking was defined as onset after 18 months. Logistic regression models of increasing complexity were fitted: (1) FC only, (2) FC plus FO, (3) FC×FO interaction, and (4) full model with sex and age at testing. Each model adds terms to the previous one (nested models). Model fit was compared using the Akaike Information Criterion (AIC; lower values indicate better model fit with fewer parameters). Model discrimination was assessed using the area under the receiver operating characteristic curve (AUC; a measure of how well the model distinguishes delayed from non-delayed children, ranging from 0.5 = chance to 1.0 = perfect) with DeLong 95% confidence intervals (95%CI, which compare two AUCs from the same sample) and the likelihood ratio test for nested model comparison. Ten-fold cross-validated AUC estimated out-of-sample performance. Sensitivity (proportion of truly delayed children correctly identified), specificity (proportion of non-delayed children correctly identified), positive predictive value (PPV; probability that a child predicted as delayed is actually delayed), and negative predictive value (NPV; probability that a child predicted as not delayed is actually not delayed) were calculated for clinical decision rules.

#### Model diagnostics

Residual normality was assessed using the Shapiro-Wilk test, heteroscedasticity using the Breusch-Pagan test, and influential observations using Cook’s distance (threshold: 4/n). Where residuals departed from normality, permutation test p-values were used to confirm parametric findings.

## Acknowledgements

The authors thank Verena Landel for her careful editorial review and constructive feedback on the manuscript. The authors also thank the children and families who participated in the vestibular assessments at the Centre d’Exploration Fonctionnelle de l’Équilibre chez l’Enfant (EFEE), Hôpital Universitaire Robert Debré (AP-HP, Paris), and the clinical staff who contributed to data collection over the study period.

## Funding

This work was supported by the European Union’s Horizon Europe research and innovation programme under the Marie Skłodowska-Curie Postdoctoral Fellowship grant agreement No. 101275781 (REHEAR) to M.C. Views and opinions expressed are those of the authors only and do not necessarily reflect those of the European Union or the European Research Executive Agency (REA). Neither the European Union nor REA can be held responsible for them. This work was further supported by a grant from Fondation Pour l’Audition (FPA) to the Institut de l’Audition (FPAIDA09 to H.T-V. and FPAIDA10 to Paul Avan and CERIAH), and by a French government grant managed by the Agence Nationale de la Recherche under the France 2030 programme (reference ANR-23-IAHU-0003). The funders had no role in study design, data collection and analysis, decision to publish, or preparation of the manuscript.

## Author contributions

M.C. designed the statistical framework, performed the analyses, interpreted the results, and wrote the manuscript. S.R.W-V. designed the study, collected the clinical data, performed vestibular assessments, and contributed to data interpretation and manuscript revision. A.M. contributed to data collection. H.T-V. contributed to manuscript revision. M.C. and S.R.W-V. contributed equally to this work. All authors approved the final manuscript.

## Competing interests

The authors declare no competing interests.

## Data availability

The clinical data supporting the findings of this study are not publicly available due to patient privacy restrictions, as they contain sensitive paediatric health information. Anonymised data are available from the corresponding author upon reasonable request and subject to appropriate data sharing agreements.

## Code availability

R code for all analyses is publicly available at https://github.com/mcampi111/Functional_Hierarchy_0tolith_Canal_Vestibular.

## Supplementary Material

**Supplementary Table 1.**
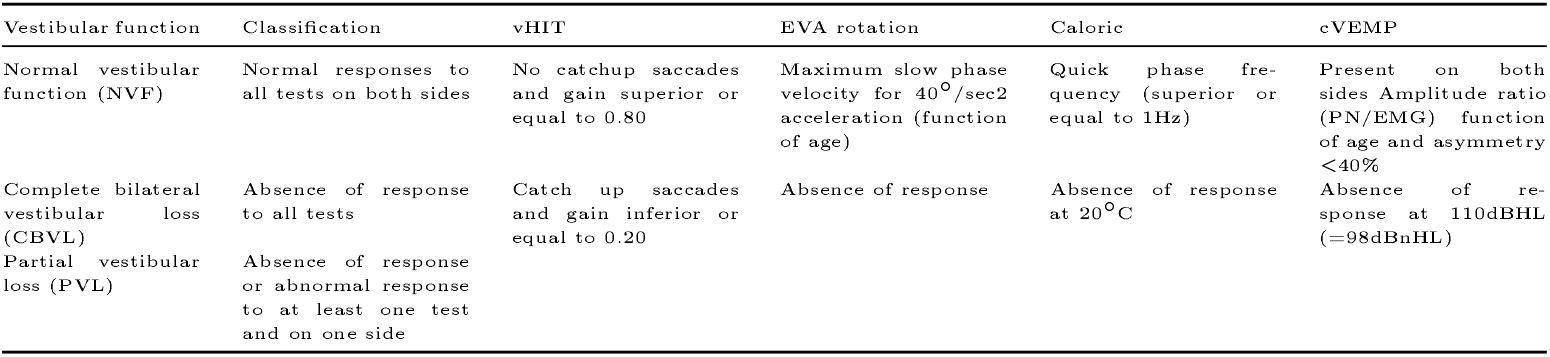
Vestibular testing criteria for classification of canal function (FC) and otolith function (FO) as normal, partial, or absent. Semicircular canal testing: video head impulse test (vHIT), earth vertical axis rotation chair test, bithermal caloric test. Otolith organ testing: cervical vestibular evoked myogenic potentials (cVEMP). NVF = normal vestibular function; CBVL = complete bilateral vestibular loss; PVL = partial vestibular loss.

| Vestibular function | Classification | vHIT | EVA rotation | Caloric | cVEMP |
| --- | --- | --- | --- | --- | --- |
| Normal vestibular function (NVF) | Normal responses to all tests on both sides | No catchup saccades and gain superior or equal to 0.80 | Maximum slow phase velocity for 40°/sec<sup>2</sup> acceleration (function of age) | Quick phase frequency (superior or equal to 1Hz) | Present on both sides Amplitude ratio (PN/EMG) function of age and asymmetry <40% |
| Complete bilateral vestibular loss (CBVL) | Absence of response to all tests | Catch up saccades and gain inferior or equal to 0.20 | Absence of response | Absence of response at 20°C | Absence of response at 110dBHL (=98dBnHL) |
| Partial vestibular loss (PVL) | Absence of response or abnormal response to at least one test and on one side |  |  |  |  |

**Supplementary Table 2.** Covariate-adjusted Type III ANOVA results (with sex and age at vestibular testing as covariates). Type III sums of squares with sum-to-zero contrasts. P-values for each factor after controlling for all other terms including sex and age at vestibular testing.

| Milestone | n | FC p | FO p | FO×FC p | Sex p | Age p |
| --- | --- | --- | --- | --- | --- | --- |
| Independent Walking | 410 | <0.001 | 0.115 | 0.222 | 0.021 | 0.437 |
| Sitting | 281 | <0.001 | 0.052 | 0.306 | 0.002 | 0.976 |
| Standing with Support | 202 | 0.157 | 0.012 | 0.071 | 0.305 | 0.351 |
| Head Holding | 197 | 0.078 | 0.273 | 0.833 | 0.614 | 0.477 |

*Note:* FC = canal function, FO = otolith function. Bold p-values indicate significance at $\alpha=0.05$ . Sex: Male=1, Female=2. Age: age at vestibular testing in months.

**Supplementary Table 3.** Graded dysfunction analysis: linear trend coefficients (FO and FC treated as continuous 0-1-2). Linear regression with FO and FC as numeric (0, 1, 2) and their interaction. Coefficients represent change in milestone age (months) per unit increase in dysfunction level.

| Milestone | FC coef (mo/level) | FC SE | FC p | FO coef (mo/level) | FO SE | FO p | Interaction coef | Interaction p | R<sup>2</sup> | Adj R<sup>2</sup> |
| --- | --- | --- | --- | --- | --- | --- | --- | --- | --- | --- |
| Independent Walking | -3.22 | 0.54 | <0.001 | -2.54 | 0.433 | <0.001 | 0.697 | 0.045 | 0.477 | 0.474 |
| Sitting | -1.4 | 0.403 | <0.001 | -0.82 | 0.36 | 0.023 | 0.181 | 0.511 | 0.27 | 0.262 |
| Standing with Support | -1.14 | 0.587 | 0.054 | -0.79 | 0.518 | 0.129 | -0.079 | 0.839 | 0.272 | 0.261 |
| Head Holding | -0.77 | 0.37 | 0.038 | -0.7 | 0.341 | 0.040 | 0.063 | 0.806 | 0.248 | 0.237 |

*Note:* Negative coefficients indicate decreasing milestone age (earlier achievement) with increasing function level (0=areflexia → 2=normal). R<sup>2</sup> and Adj R<sup>2</sup> for the full model including FO, FC, and interaction.

**Supplementary Table 4.** Bootstrap confidence intervals for adjusted delays and canal-to-otolith ratios (5,000 BCa resamples). Bias-corrected and accelerated (BCa) bootstrap 95% CIs. Standing excluded due to significant interaction. Delays are estimated marginal means contrasts (areflexia vs normal).

| Milestone | Canal delay (mo) | Canal CI low | Canal CI high | Otolith delay (mo) | Otolith CI low | Otolith CI high | Ratio | Ratio CI low | Ratio CI high |
| --- | --- | --- | --- | --- | --- | --- | --- | --- | --- |
| Independent Walking | 7 | 3.9 | 9.2 | 2.2 | 0.9 | 3.4 | 3.8 | 1.5 | 8.7 |
| Sitting | 2.2 | -0.8 | 5.4 | 0.7 | 0 | 1.9 | 7.7 | -0.9 | 41.3 |
| Head Holding | 1.9 | 0.7 | 2.9 | 0.8 | -0.1 | 1.7 | 8.9 | 0.6 | 34.1 |

*Note:* Ratio = canal delay ÷ otolith delay. Wide ratio CIs reflect small otolith denominators. BCa method corrects for bias and skewness in the bootstrap distribution.

**Supplementary Table 5.** Permutation test results (10,000 permutations). Non-parametric significance tests. FC and FO labels permuted independently; p = proportion of permuted F-statistics ≥ observed F-statistic.

| Milestone | FC F-observed | FC permutation p | FO F-observed | FO permutation p |
| --- | --- | --- | --- | --- |
| Independent Walking | 16.8 | <0.001 | 2.57 | 0.0764 |
| Sitting | 6.52 | 0.0035 | 3.63 | 0.0303 |
| Standing with Support | 1.98 | 0.1417 | 5.43 | 0.0059 |
| Head Holding | 2.33 | 0.0993 | 1.44 | 0.2309 |

*Note:* Permutation tests make no distributional assumptions. Results largely confirm parametric Type III ANOVA findings. FC walking $p < 0.001$ in both approaches; FO walking $p = 0.076$ (permutation) vs $p = 0.077$ (ANOVA).

**Supplementary Table 6.** Age-stratified validation (children ≤24 months at vestibular testing, n=270). Replication of primary analysis in younger subsample to assess robustness. Same Type III ANOVA approach; main-effects-only model used when empty cells preclude interaction estimation.

| Milestone | n | Canal delay (mo) | Canal p | Otolith delay (mo) | Otolith p | Ratio | Interaction p | Note |
| --- | --- | --- | --- | --- | --- | --- | --- | --- |
| Independent Walking | 270 | 7.6 | <0.001 | 3 | 0.024 | 2.54 | 0.101 |  |
| Sitting | 196 | 4.5 | <0.001 | 0.8 | 0.392 | 5.86 | 0.168 |  |
| Standing with Support | 142 | 3.2 | <0.001 | 2 | 0.008 | 1.58 | — | Main effects only (empty cells) |
| Head Holding | 141 | 1.5 | 0.180 | 1.1 | 0.184 | 1.38 | 0.998 |  |

*Note:* Canal dominance for walking replicated in younger subsample (7.6 mo delay, $p < 0.001$ ). Standing uses main-effects-only model due to empty FO × FC cell (FO=1, FC=2: $n = 0$ in $\leq 24$ mo subsample).

**Supplementary Table 7.** Robust sample validation (FO×FC combinations with n≥10 only, n=395). Excludes combinations with small sample sizes (FO=0/FC=2, FO=2/FC=0, FO=1/FC=2: all n<5) to assess whether results are driven by small-sample combinations. Main-effects-only model used due to empty cells after exclusion.

| Milestone | n | Canal delay (mo) | Canal p | Otolith delay (mo) | Otolith p | Ratio | Interaction p | Note |
| --- | --- | --- | --- | --- | --- | --- | --- | --- |
| Independent Walking | 394 | 4.3 | <0.001 | 4.2 | <0.001 | 1.02 | — | Main effects only (empty cells) |
| Sitting | 266 | 2 | 0.001 | 1.6 | 0.001 | 1.24 | — | Main effects only (empty cells) |
| Standing with Support | 195 | 2.6 | 0.004 | 1.6 | 0.031 | 1.62 | — | Main effects only (empty cells) |
| Head Holding | 189 | 1.2 | 0.047 | 1.4 | 0.005 | 0.86 | — | Main effects only (empty cells) |

*Note:* In the robust sample, canal and otolith delays converge (ratio $\approx 1.0$ for walking), suggesting that the canal dominance in the full sample partly reflects the confounding structure of combinations with small sample sizes. All models use main effects only due to empty cells after exclusion.

**Supplementary Table 8.** Model diagnostics for Type III ANOVA models. Normality (Shapiro-Wilk), homoscedasticity (Breusch-Pagan), and influential observations (Cook’s distance > 4/n) for the FO×FC interaction model on each milestone.

| Milestone | Shapiro-Wilk W | Shapiro-Wilk p | Breusch-Pagan stat | Breusch-Pagan p | N influential | N total |
| --- | --- | --- | --- | --- | --- | --- |
| Independent Walking | 0.9666 | <0.001 | 24.13 | 0.0022 | 21 | 410 |
| Sitting | 0.9268 | <0.001 | 33.46 | <0.001 | 17 | 281 |
| Standing with Support | 0.9675 | <0.001 | 17.54 | 0.0250 | — | 202 |
| Head Holding | 0.9362 | <0.001 | 25.92 | 0.0011 | — | 197 |

*Note:* All milestones show significant non-normality (Shapiro-Wilk $p < 0.001$ ) and heteroscedasticity (Breusch-Pagan $p < 0.05$ ). This motivates the permutation tests (Suppl. Table 4) and bootstrap CIs (Suppl. Table 3) as non-parametric sensitivity analyses. ANOVA is robust to moderate violations with $n > 200$ .

**Supplementary Table 9.** Ten-fold cross-validated classification performance for walking delay prediction (>18 months). Logistic regression models evaluated by 10-fold CV. Threshold optimised for each fold. Performance averaged across folds.

| Model | CV-AUC | Sensitivity | Specificity | PPV | NPV |
| --- | --- | --- | --- | --- | --- |
| Canal only | 0.796 | 0.566 | 0.936 | 0.802 | 0.824 |
| Otolith only | 0.769 | 0.744 | 0.776 | 0.604 | 0.869 |
| Canal + Otolith | 0.831 | 0.55 | 0.911 | 0.74 | 0.815 |
| Full | 0.852 | 0.581 | 0.89 | 0.708 | 0.822 |

*Note:* CV-AUC = cross-validated area under the ROC curve. PPV = positive predictive value. NPV = negative predictive value. Canal+Otolith model (CV-AUC=0.831) outperforms Canal only (0.796) and Otolith only (0.769). Full model with interaction achieves highest CV-AUC (0.852) but marginal improvement over additive model.

## Notes

### Competing Interest Statement

The authors have declared no competing interest.

### Author Declarations

Ethics committee CPP-Ouest 3 of Hopital Universitaire Robert Debre AP-HP, Paris gave ethical approval for this work (reference 2018-A01705-50). Written consent requirements were waived given the retrospective nature of the study.

